# EARLY VIRAL CLEARANCE AMONG COVID-19 PATIENTS WHEN GARGLING WITH POVIDONE-IODINE AND ESSENTIAL OILS – A CLINICAL TRIAL

**DOI:** 10.1101/2020.09.07.20180448

**Authors:** Nurul Azmawati Mohamed, Nizam Baharom, Wan Shahida Wan Sulaiman, Zetti Zainol Rashid, Wong Kon Ken, Umi Kalsom Ali, Siti Norlia Othman, Muttaqillah Najihan Samat, Najma Kori, Petrick Periyasamy, Nor Azizan Zakaria, Agni Nhirmal Kumar Sugurmar, Nur Ezzaty Mohammad Kazmin, Cheong Xiong Khee, Siti Mariyam Saniman, Ilina Isahak

## Abstract

**Background:** Gargling had been reported to have significant roles in the prevention and treatment of respiratory tract infections. The purpose of this study was to assess the ability of regular gargling to eliminate SARS-CoV-2 in the oropharynx and nasopharynx.

**Methodology:** This pilot, open labeled, randomized, parallel study compared the effect of 30 seconds, 3 times/day gargling using 1% povidone-iodine (PVP-I), essential oils and tap water on SARS-CoV-2 viral clearance among COVID-19 patients in a tertiary hospital in Kuala Lumpur. Progress was monitored by day 4,6 and 12 PCR (Ct value), gargling and symptoms diary as well as clinical observations.

**Results:** Five confirmed Stage 1 COVID-19 patients were recruited for each arm. The age range was from 22 to 56 years old. The majority were males. Two respondents had co-morbidities, which were asthma and obesity. Viral clearance was achieved at day 6 in 100%, 80%, 20% and 0% for 1% PVP-I, essential oils, tap water and control group respectively. Analysis of 1% PVP-I group versus control group showed significant p-value for comparison of PCR results on Day 4, Day 6 and Day 12.

**Conclusions:** This preliminary study showed that gargling with 1% PVP-I and essential oils show great potential to be part of the treatment and management of Stage 1 COVID-19. Larger studies are required to ascertain the benefit of gargling for different stages of COVID-19 patients. This study was registered in clinicaltrial.gov (NCT04410159).

## Introduction

Coronavirus disease 2019 (COVID-19) that is caused by SARS-CoV-19 was first recognized in December (Du Toit 2020). As of 9^th^ July 2020, the disease has spread rapidly to more than 200 countries, infected more than 12 million people and caused more than 500,000 deaths worldwide. The virus mainly transmits via respiratory droplets when infected persons sneeze or cough and through contaminated surfaces. The R_0_ of SARS-CoV-2 was estimated to be 3.58 (from person to person), which means that the expected number of secondary infections resulted from introducing a single infected individual into a susceptible population was 3.58 (Chen et al. 2020). To date, there is neither licensed medication nor vaccine to treat and prevent COVID-19. The management of the disease relies on symptomatic treatment and outbreak preventive measures such as hand hygiene, social distancing and wearing a mask. Therefore, there is an intense need to explore new management strategies to treat and prevent the disease.

The course of disease for COVID 19 is divided into 3 stages (Mason 2020). Stage 1 is an asymptomatic state start at the beginning of the first two days of infection. Stage 2 followed as the disease progresses with the development of mild symptoms such as fever and cough and the third stage started when the virus infiltrates the lungs with the manifestation of more severe symptoms. The establishment of disease begins with the entry of the virus into the upper respiratory tract (nasal cavity and pharynx) and eyes where the virus adheres to the mucosa epithelium and primary replication believed to occur here before it further replicates in the lung (Jin et al. 2020). As the viral load was found highest in the nasopharynx, high in saliva and throat, these areas likely seed the lower respiratory tract and serve as the main reservoirs for droplet transmission and pulmonary disease progression (Mady et al. 2020). Considering the pathogenesis of COVID-19, there is a role of gargle solution with virucidal property in achieving viral clearance or interrupting the disease progression (Pattanshetty, Narayana, and Radhakrishnan 2020). Viral clearance is defined as the presence of negative SARS-COV-2 polymerase chain reaction (PCR) results from at least two upper respiratory tract samples, collected at ≥ 24-hour intervals (ECDC 2020).

Gargling involves washing of the oral cavity and pharynx with a liquid gargle. During the gargling process, mucus will loosen and rinse irritants such as allergens, and pathogens inside the oral cavity including part of the oropharynx region. Gargling is also believed to bring about favorable effects through the removal of oral/pharyngeal protease that helps viral replication (Kitamura et al. 2007). There are studies that have been done reported on the effectiveness of gargling with various solutions as prevention or treatment of respiratory tract infection from as simple as water gargling to the use of povidone-iodine, chlorhexidine, green tea and essential oil-based formula (Satomura et al. 2005; Kitamura et al. 2007; Toyoizumi et al. 2013).

A randomized controlled trial in community healthcare settings showed that simple water gargling has significantly reduced the incidence rate of URTIs by 36%. This result was explained by two possibilities: whirling water washed out pathogens from the pharynx and oral cavity and the presence of high chlorine in the tap water might deactivate viruses (Satomura et al. 2005). Another study reported that the cumulative incidence of URTI and influenza among school children was reduced by 24% when gargling with diluted 7% povidone-iodine (PVP-I) thrice daily (Kitamura et al. 2007). The effectiveness of green tea gargle in reducing influenza had also been tested. However, the result showed no significant difference when compared to the control group (Toyoizumi et al. 2013).

Challacombee *et al* proposed a protocolised intra-nasal and oral application of PVP-I for both patients and their attendant healthcare workers (HCWs) during the current COVID-19 pandemic to help limit the spread of SARS-CoV-2 from patients to healthcare workers and vice versa (Cascella et al. 2020) (Challacombe et al. 2020). To date, there is no published report on the effectiveness of gargling in limiting the spread of SARS-CoV-2. Therefore, we embarked on this preliminary study to look for the effect of gargling with PVP-I, essential oils, and tap water among COVID-19 patients on viral clearance. 1% PVP-I was chosen due to its proven virucidal activity against MERS-CoV and SAR-CoV (Eggers et al. 2018). However, the formulation is not widely available and contraindicated in those with thyroid abnormality. Hence, we also tested an essential oil formula that is widely available in the market and will serve as an alternative for those with thyroid disorders.

## Material and Methods

### Study design

This 4-arms preliminary interventional study compared the effect of gargling with 1% PVP-I (Betadine®), essential oils (Listerine® Original) and tap water among Stage 1 COVID-19 patients. A control group with no intervention was also included in the study.

### Sample size and population

For this pilot study, we looked at Eggers et al. (2015), using Betadine® gargle on MERS-CoV which showed a significant reduction of viral titer by a factor of 4.3 log_10_ TCID50/mL, and we calculated a sample size of 5 per arm or 20 samples in total. This study involved 20 COVID-19 patients from one university hospital with designated COVID-19 center. Inclusion criteria included adults aged 18 years and above, asymptomatic COVID-19 patients (Stage 1) and less than 5 days from diagnosis (first positive swab sample). Meanwhile, exclusion criteria were; unable to understand instructions, those with any respiratory symptoms or fever on admission, abnormal chest radiograph or computed tomography (CT) findings on admission, those started on treatment for COVID-19 or COVID-19 complication (e.g enoxaparine sodium (clexane)), those reinfected with SARS-CoV-2, abnormal thyroid function test and allergy to PVP-I (both for group A). Patients were randomly assigned using 20 identical envelopes, each containing a card nominating either one of the four arms; gargle with 1% PVP-I (Group A), gargle with essential oils (Group B), gargle with tap water (Group C) and no intervention (Group D). This study was open labelled and there was no masking or stratification done. All patients were recruited on the day of being diagnosed with COVID-19 and commenced gargle activity on the day after.

### Experimental plan

After consent was taken, all groups were briefed regarding the study protocol separately.

- Group A patients were briefed on the correct procedures of gargling with Betadine®. The patients were instructed to take 10ml of Betadine® gargle/mouthwash, tilt their heads backward and gargle for 30 seconds, three times per day for 7 days.
- Group B patients were briefed on the correct technique of gargling with Listerine®. The patients were instructed to take 20ml of Listerine®, tilt their heads backward and gargle for 30 seconds, three times per day for 7 days
- Group C patients were briefed on the correct technique of gargling with tap water. The patients were instructed to take 100mL of tap water, tilt their heads backward and gargle for 30 seconds, three times per day for 7 days
- Group D patients were briefed about the involvement in this study. They were managed according to the standard protocol of the hospital with no additional intervention.

### Monitoring

Patients were given a chart for them to write down the frequency of gargling and symptoms (if any). Clinical and laboratory progression were monitored for 7 days as shown in the diagram below. Nasopharyngeal and oropharyngeal swabs were taken at day 4,6 and 12 of the intervention. The swabs were taken before the early morning gargle. Vital signs, chest radiograph findings and absolute lymphocytic counts (ALC) were recorded in the clinical data collection sheet.

### Detection of SAR-CoV-2 by real time RT-PCR

Viral RNAs were extracted from nasopharyngeal and oropharyngeal samples using GeneAll® GENTi™ Viral DNA/RNA Extraction Kit according to the manufacturer’s manual. Real time reverse transcription polymerase chain reaction (RT-PCR) was performed using a commercial kit, LyteStar™ 2019-nCoV RT-PCR Kit 1.0. The kit consisted of two independent assays, targeting the *E* gene that detected subgenus Betacoronavirus and RNA-dependent RNA polymerase gene *(RdRP)* that was specific for the SARS-CoV-2 genome. In both assays, probes specific for the *E* gene and *RdRP* gene were labeled with fluorophore FAM™.

Each RT-PCR assay was provided with a Ct (cycle threshold) value. The cycle threshold is the number of cycles required for the fluorescent signal to cross the threshold (exceeds the background level). It is inversely correlated with viral load. The specimens were considered positive if the Ct value for both assays were 45.0 or lower and negative results referred to no Ct obtained. Indeterminate results were reported when only one gene assay showed Ct of less than 45.0.

### Data Analysis

Statistical analysis was performed using the IBM SPSS Statistics (v24.0) for descriptive and inferential statistics, such as the Fisher-Freeman-Halton exact test to determine associations between gargling groups and COVID-19 swab results.

### Ethical Approval

This study received approval from the Research and Ethics Committee, Faculty of Medicine, Universiti Kebangsaan Malaysia (UKM PPI/111/8/JEP-2020-372). This study was registered in clinicaltrial.gov (NCT04410159).

## Results

Twenty COVID-19 positive patients participated in the study (Table 1). The youngest and oldest respondents were 22 and 56-year-old, respectively. The majority were males and all patients were asymptomatic and classified as stage 1 of COVID-19. Two respondents had co-morbidities, which were asthma and obesity. There were no abnormal vital sign changes, full blood count or chest X-ray findings from any of the patients.

**Table 1:**
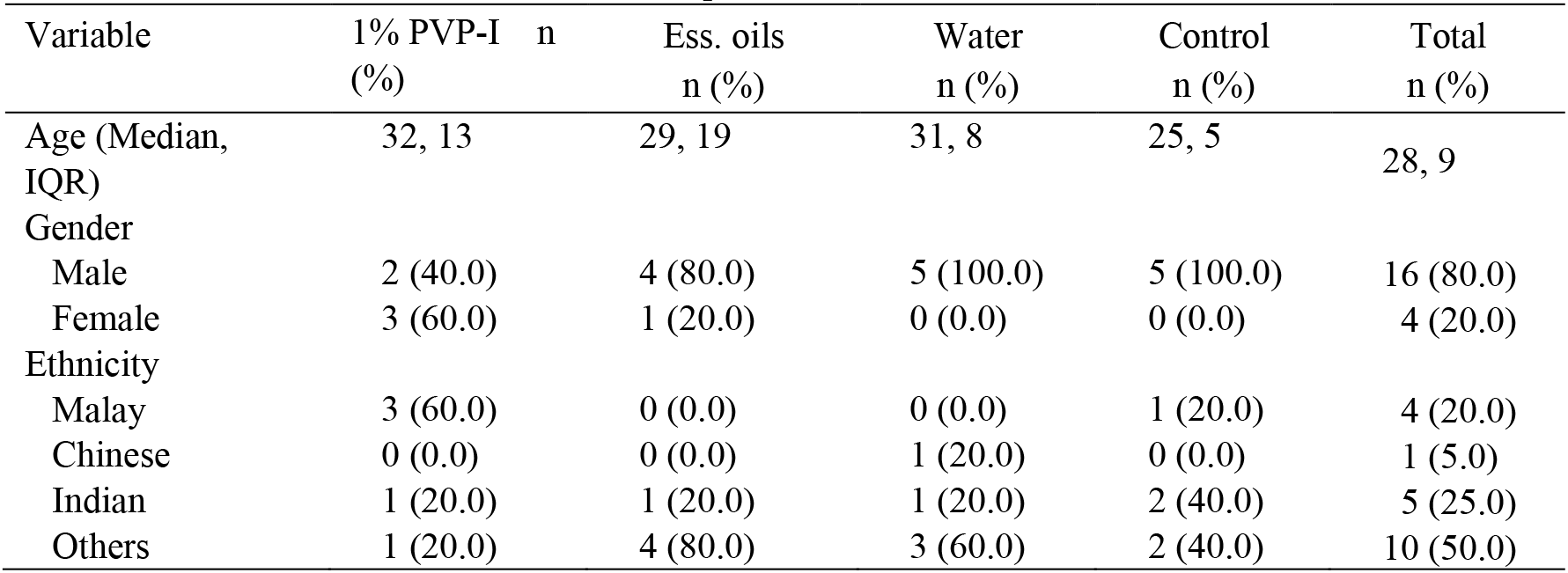
Socio-clinical characteristics of the respondents, n=20

Among all 5 respondents in the 1% PVP-I group, SARS-CoV-2 was not detected on Day 4, Day 6 and Day 12 (Table 2). Within the essential oils group, there were 4 negative samples with 1 either positive or indeterminate result throughout Day 4, Day 6 and Day 12 swab tests. Within the tap water group, 2 remained negative while the other 3 were either positive or indeterminate for SARS-CoV-2 on Day 4, Day 6 and Day 12. Within the control group, only one sample was negative on Day 4 and Day 12, while all other samples were either positive or indeterminate. Viral clearance was achieved in 100%, 80%, 20% and 0% for 1% PVP-I, essential oils, tap water gargle and control group respectively. There was no reporting of any side effects from the use of 1% PVP-I, essential oils or tap water.

**Table 2:** COVID-19 PCR results for swabs at day 4, 6 and day 12 post-intervention

| Group | Day 4, n (%) |  |  | Day 6, n (%) |  |  | Day 12, n (%) |  |  |
| --- | --- | --- | --- | --- | --- | --- | --- | --- | --- |
|  | Neg | Pos | Ind | Neg | Pos | Ind | Neg | Pos | Ind |
| 1% PVP-I,<br>n=5 | 5<br>(100.0) | 0<br>(0.0) | 0<br>(0.0) | 5<br>(100.0) | 0<br>(0.0) | 0<br>(0.0) | 5<br>(100.0) | 0<br>(0.0) | 0<br>(0.0) |
| Essential oils, n=5 | 4<br>(80.0) | 1<br>(20.0) | 0<br>(0.0) | 4<br>(80.0) | 1<br>(20.0) | 0<br>(0.0) | 4<br>(80.0) | 0<br>(0.0) | 1<br>(20.0) |
| Tap Water, n=5 | 2<br>(40.0) | 3<br>(60.0) | 0<br>(0.0) | 2<br>(40.0) | 1<br>(20.0) | 2<br>(40.0) | 2<br>(40.0) | 2<br>(40.0) | 1<br>(20.0) |
| Control, n=5 | 1<br>(20.0) | 2<br>(40.0) | 2<br>(40.0) | 0<br>(0.0) | 3<br>(60.0) | 2<br>(40.0) | 1<br>(20.0) | 3<br>(60.0) | 1<br>(20.0) |

Neg = Negative, Pos = Positive, Ind = Indeterminate result.

Table 3 shows the analysis of the 1% PVP-I group versus the control group using the Fisher-Freeman-Halton exact test. The *p* values were significant for comparison of samples results on Day 4, Day 6 and Day 12. A similar analysis for 1% PVP-I versus tap water group, Listerine versus the control group and Listerine versus tap water group were all not significant (results not shown).

**Table 3:**
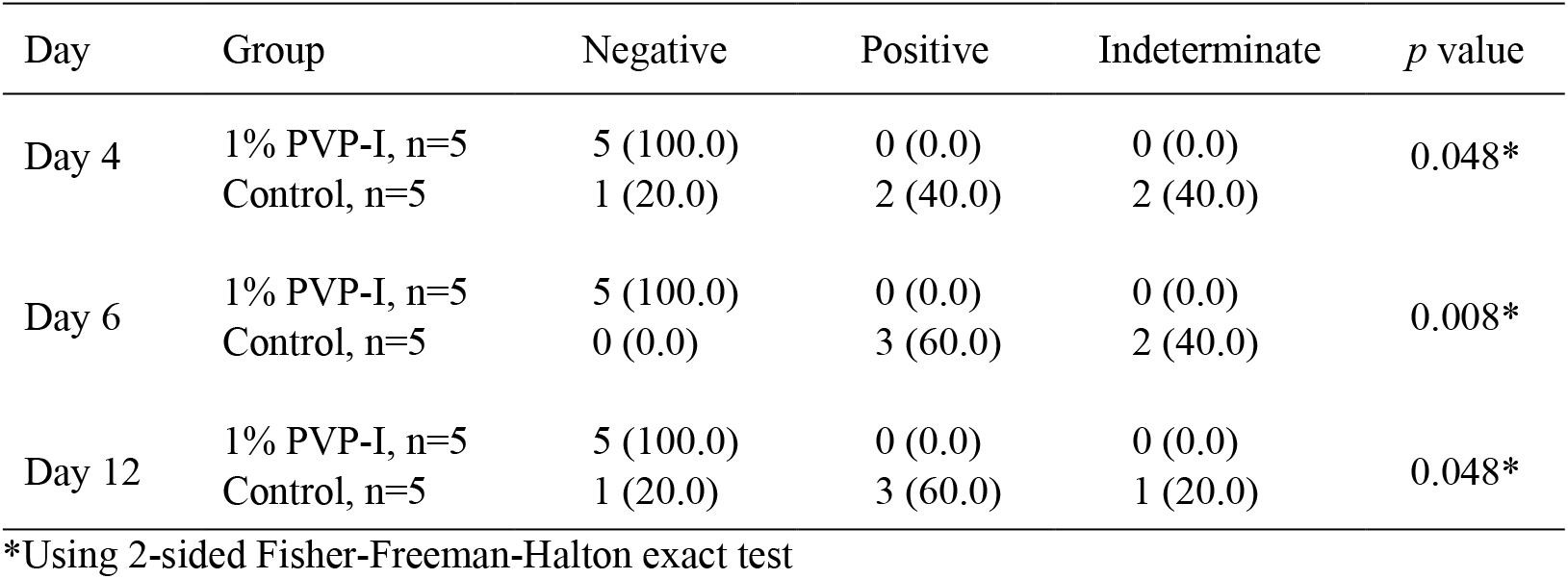
Comparing the COVID-19 PCR results of 1% PVP-I group versus control at Day 4, 6 and 12.

| Day | Group | Negative | Positive | Indeterminate | *p* value |
| --- | --- | --- | --- | --- | --- |
| Day 4 | 1% PVP-I, n=5 | 5 (100.0) | 0 (0.0) | 0 (0.0) | 0.048* |
|  | Control, n=5 | 1 (20.0) | 2 (40.0) | 2 (40.0) |  |
| Day 6 | 1% PVP-I, n=5 | 5 (100.0) | 0 (0.0) | 0 (0.0) | 0.008* |
|  | Control, n=5 | 0 (0.0) | 3 (60.0) | 2 (40.0) |  |
| Day 12 | 1% PVP-I, n=5 | 5 (100.0) | 0 (0.0) | 0 (0.0) | 0.048* |
|  | Control, n=5 | 1 (20.0) | 3 (60.0) | 1 (20.0) |  |

\*Using 2-sided Fisher-Freeman-Halton exact test

## Discussions

It has been shown that without intervention, median time from diagnosis to viral clearance was approximately two weeks for asymptomatic patients (Uhm et al. 2020). However, this current study showed high viral clearance rate for 1% PVP-I and essential oils gargles as early as 4 days after intervention (equivalent to 5 to 6 days after diagnosis). Early viral clearance is very critical in COVID-19 as the disease is highly infectious at the early stage. A study found pharyngeal viral shedding was highest at day 4 of presentation, and successful isolation of the virus suggested active viral replication (Wölfel et al. 2020).

The viral clearance could be explained by the use of active virucidal ingredients and the mechanical effect of gargling. A recent study demonstrated that 1% PVP-I achieved >5 logs 10 reductions in the SARS CoV-2 virus titre at 15, 30 and 60 seconds (Hassandarvish et al. 2020). Is was also proven effective against Severe Acute Respiratory Syndrome coronavirus (SARS-CoV) and the Middle East Respiratory Syndrome coronavirus (MERS-CoV) (Eggers et al. 2018; Eggers, Eickmann, and Zorn 2015). PVP-I is one of the antiseptics widely used in the clinical setting. It has multiple microbicidal mechanisms of actions including inhibition of vital bacterial cellular mechanisms and structures, oxidization of nucleotides fatty/amino acid and cytosolic enzymes (Kanagalingam et al. 2015). In contrast to other antiseptics such as chlorhexidine and quaternary ammonium salts, no acquired resistance or cross-resistance has been reported for iodine in over 150 years of use (Bigliardi et al. 2017).

Commercial essential oils for mouthwash and gargle contains a combination of four types of essential oils namely thymol, eucalyptol, methyl salicylate and menthol. Generally, essential oils disrupt viral membrane or interfere with viral envelope protein involved in host cell attachment (Nadjib 2020). A study showed that exposure to widely used essential oil formulas i.e. Listerine® had an antiviral effect against herpes simplex virus type 1 (Meiller et al. 2005). Eucalyptus has been shown to directly inactivate free virus particles and interfere with virion envelope structures of herpes simplex 1 (Astani, Reichling, and Schnitzler 2010). Even though essential oils have not been tested against coronaviruses, a similar effect on viral membrane or envelope might be observed for SARS-CoV-2. This may explain the high viral clearance rate among the essential oils group in this study.

Asymptomatic Stage 1 patients can be considered as a source of COVID-19 and able to transmit the virus to close contacts. Some of the infected close contacts even had very severe pneumonia and required intensive care monitoring (Hu et al. 2020). The findings of this preliminary study suggest that regular gargling with 1% PVP-I or essential oils formula for Stage 1 or early stage of COVID-19 patients has great potential to minimize viral transmission. This is in line with a systematic review that showed the incidence of pneumonia could be reduced by oral hygiene measures using chlorhexidine and PVP-I (Manger et al. 2017).

Gargling also seems to have the potential to reduce the disease spread from patients to healthcare workers. This is in agreement with Mady et al who described a gargling protocol for healthcare workers caring for head and neck and skull base oncology patients (Mady et al. 2020). Spread among the community could also be reduced with gargling practices among the public after being exposed to crowded places or high-risk areas. Nevertheless, this measure is not to replace but to complement currently practiced preventive measures namely hand washing, mask-wearing and social distancing.

This study was limited by several factors. Firstly, a larger sample size would be ideal for this pilot. However, we were limited by dwindling numbers of COVID-19 patients. Secondly, frequency, duration and method of gargling could only be monitored by a daily chart that was filled by patients themselves. For example, one patient from the essential oils group did not achieve viral clearance at day 6. The respondent’s PCR was only tested negative at day 17 after the intervention, similar to those in the control group. This condition could be attributed to low compliance, poor gargling technique or lack of response to the essential oil gargle. Thirdly, baseline (pre-intervention) Ct value that indicated initial viral load was not available for comparison with subsequent Ct values. Knowing the baseline value would be beneficial in comparing the effectiveness of gargling among those with higher and lower viral load. Finally, the actual day of viral clearance might be shorter; however, the PCR test was not done before day 4.

## Conclusions

This preliminary study showed that regular gargling with 1% PVP-I and essential oils formula have the potential for achieving early SARS-CoV-2 viral clearance among stage 1 COVID-19 patients. Larger studies are required to ascertain the benefits of gargling for prevention and management of different stages of COVID-19 patients.

## Data Availability

Data is available upon request

https://clinicaltrials.gov/ct2/show/NCT04410159?cond=gargle+covid&draw=2&rank=7

## Acknowledgement

We would like the thank the Director of the Chancellor Tuanku Mukhriz Hospital and the Research Committee of Faculty of Medicine, Universiti Kebangsaan Malaysia for the permission granted for this study. This study was self-funded by the research team.

## Author’s Contribution

NAM and NK were the co-principal investigator, NB and WSWS designed the study and analysed the data, PP and NAZ supervised the study implementation, NEMK, CXK, ANKS and SMS monitored participants and collected data, UKA, SNO and WKK involved in molecular investigations, ZZR and MNAS wrote the draft, II supervised the study and proofread the manuscript.

## Declaration

not applicable

## Availability of data

upon request

